# Leisure-time physical activity reduces the risks of mortality associated with general and abdominal obesity in adults in Mexico

**DOI:** 10.1101/2025.05.30.25328656

**Authors:** Gary O’Donovan, Fanny Petermann-Rocha, Gerson Ferrari, Catalina Medina, Evelia Apolinar-Jiménez, Olga L. Sarmiento

**Author notes:** Corresponding author (GOD).

## Abstract

**Aim:** Obesity is one of the leading public health problems in Mexico. Therefore, we investigated whether physical activity reduced the risks of mortality associated with obesity in participants in the Mexico City Prospective Study.

**Methods:** A total of 153,060 adults aged 52 (13) years were followed for 18 (4) years [mean (SD)]. Trained nurses asked about leisure-time physical activity and measured height, weight, and waist circumference. Cox models were adjusted for confounders and cardiovascular disease (CVD) and cancer mortality were treated as competing risks.

**Results:** There were 25,317 deaths from all causes, 8,488 from CVD, and 3,368 from cancer during follow-up. Compared with the group that reported little or no activity and had a normal body mass index, the hazard ratio (95 % confidence interval) for all-cause mortality was 1.40 (1.33, 1.47) in the group that reported little or no activity and had Obesity Class II. Physical activity reduced the risk of all-cause mortality associated with obesity. For example, the hazard ratio was 1.18 (1.06, 1.32) in the group that reported medium or high activity and had Obesity Class II. Physical activity also reduced the risk of CVD mortality associated with Obesity Class II and the risk of cancer mortality associated with Obesity Class I and Obesity Class II. Furthermore, physical activity reduced the risks of all-cause, CVD, and cancer mortality associated with high waist circumference.

**Conclusions:** This novel study suggests that leisure-time physical activity reduces the risks of mortality associated with general and abdominal obesity in adults in Mexico.

## Introduction

Obesity has recently been described as the leading public health problem in Mexico.^1^ Data from the Global Burden of Disease Study suggest that obesity, high fasting glucose, and high blood pressure are the leading causes of morbidity and mortality across Latin America.^2^ However, the available evidence suggests that the burden of obesity is particularly great in Mexico.^3^ Therefore, it is concerning that the prevalence of obesity is continuing to rise every year in Mexico.^4^ Weight loss is incredibly difficult at the population level^5, 6^ and it has been suggested that the promotion of healthy lifestyles is a more effective strategy than the promotion of weight loss in reducing the burden of obesity.^6, 7^ Indeed, cohort studies in Europe and North America suggest that a physically active lifestyle can reduce the risks of mortality associated with general and abdominal obesity.^8–10^ The results of cohort studies in Europe and North America cannot be generalized to Latin America because of the great differences in income, health, and healthcare between the regions.^11–14^ Therefore, the main objective of the present analysis was to investigate whether leisure-time physical activity reduced the risks of mortality associated with general and abdominal obesity in more than 150,000 participants in the Mexico City Prospective Study.

## Methods

### Participants

The Mexico City Prospective Study is described in detail elsewhere.^15^ Briefly, door-to-door interviews were conducted from 1995 to 1997 to compile a record of all households in the neighboring districts of Coyoacán and Iztapalapa in Mexico City in Mexico.^15^ Recruitment teams then visited households and at least one participant aged 35 years or older was recruited from 94% of eligible households.^15^ The resulting sample was deemed to be broadly representative of the population aged 35 years or older in Mexico City.^15^ The baseline survey took place from 1998 to 2004 and mortality was tracked to 31^st^ December 2020. The study was approved by the Mexican Ministry of Health, the Mexican National Council of Science and Technology (approval number 0595 P-M), and the Central Oxford Research Ethics Committee (approval number C99.260). Trained nurses collected data in the participant’s household and all participants provided written informed consent. The data used in the current report were obtained through an open-access data request made to the Mexico City Prospective Study principal investigators.

### Exposures

The exposures were leisure-time physical activity, body mass index (BMI), and waist circumference at baseline. Leisure-time physical activity is a broad term that includes sport and exercise.^16–18^ Leisure-time physical activity was assessed using three simple questions. Participants were asked whether they exercised or played sports (yes or no). Those who said yes, were then asked how many times per week they exercised (less than once per week; once or twice per week; or, three or more times per week). Finally, they were asked how many minutes they spent exercising per session (less than 30; from 30 to 60; or, more than 60). Leisure-time physical activity volume was categorized as none or low or, alternatively, medium or high as described in Supplemental Table S1 in the online supplement. Simple physical activity assessment tools like the one used in the present study have been validated against multiple-item physical activity assessment tools^19^ and against cardiorespiratory fitness.^20^ The nurses measured weight and height in light clothing and without shoes and we expressed BMI as weight in kilograms divided by height in meters squared. Normal BMI was defined as 18.5-24.9 kg/m^2^, underweight as <18.5 kg/m^2^, overweight as 25.0-29.9 kg/m^2^, Obesity Class I as 30.0-34.9 kg/m^2^, and Obesity Class II as ≥35 kg/m^2^.^21^ The nurses measured waist circumference at the mid-point between the iliac crest and the costal margin using an inelastic tape. In the main analysis, low waist circumference was defined as <88 cm in women and <102 cm in men while high waist circumference was defined as ≥88 cm in women and ≥102 cm in men.^21^

### Outcomes

The registration of deaths in Mexico is thorough, with almost all deaths certified medically and with few deaths attributed to unknown causes.^22^ The outcomes were all-cause mortality, CVD mortality, and cancer mortality. Mortality was tracked through probabilistic linkage to the national death register based on the participant’s name, age, and sex. Diseases listed on the death certificates were coded according to the International Classification of Diseases, 10^th^ Revision (ICD-10).^23^

### Potential confounders

Potential confounders that may influence the relationships between physical activity, adiposity, and mortality include age, sex, education, income, civil status, smoking, alcohol, diet, and sleep.^8, 24–26^ The nurses recorded age and sex. Participants were asked about education and we created five groups (none; at least some elementary; at least some high-school; at least some college; and, at least some university). Participants were asked about income and we created three categories (tertile 1, tertile 2, and tertile 3). Participants were asked about their civil status and we created three groups (not married or with partner; married or with partner; widowed). The nurses asked about smoking and we created three categories (never, former, and current). Participants were also asked about alcohol drinking frequency and we created four categories (never; occasionally; at least once per week; and, daily). Diet quality was expressed as fruit and vegetable intake (never; one or two days per week; three or four days per week; five or more days per week). Finally, participants were asked about sleep duration and the at-risk group was defined as sleeping less than seven or more than nine hours per night.

### Statistical analyses

Participants’ characteristics at baseline were described according to leisure-time physical activity volume. In the main analyses, we investigated associations of physical activity, BMI, and waist circumference at baseline with all-cause, CVD, and cancer mortality at follow-up. Cox models were created and hazard ratios and 95 % confidence intervals were calculated. The proportional hazards assumption was checked graphically for the discrete physical activity volumes and no violations were observed. For CVD mortality, all other causes of death were treated as competing risks. For cancer mortality, all non-cancer deaths were treated as competing risks. Competing risks were estimated using the methods described by Fine and Gray.^27^ We first investigated the independent associations of leisure-time physical activity volume, BMI, and waist circumference with mortality. Model 1 was adjusted for age and sex. Model 2 was adjusted for age and sex; additionally, the physical activity model was adjusted for BMI (continuous variable) and waist circumference (continuous variable), the BMI model was adjusted for waist circumference (continuous variable), and the waist circumference model was adjusted for BMI (continuous variable). Model 3 was adjusted for age, sex, education, income, civil status, smoking, alcohol, fruit and vegetable intake, and sleep. We then investigated the combined associations of leisure-time physical activity volume and BMI with mortality after adjusting for age, sex, education, income, civil status, smoking, alcohol, fruit and vegetable intake, and sleep. Participants with underweight were not included in the analysis because of the extremely low number of cohort members with the condition. Finally, we investigated the combined associations of leisure-time physical activity volume and waist circumference with mortality after adjusting for age, sex, education, income, civil status, smoking, alcohol, fruit and vegetable intake, and sleep. In the sensitivity analyses we defined abdominal obesity as waist circumference ≥80 cm in women and ≥90 cm in men because these are the definitions typically used in adults in Mexico.^4^ Data from the first two years of follow-up were not used to minimize the possibility of reverse causation. All analyses were performed using Stata MP version 18.0 for Mac (StataCorp, Texas, USA).

## Results

Data from 153,060 of 159,517 (96 %) cohort members were used in the present analysis. Table 1 shows participants’ characteristics at baseline according to leisure-time physical activity volume. The average age was 52 years in those who reported little or no leisure-time physical activity and in those who reported medium or high leisure-time physical activity. The proportion of women was 70 % in the group that reported little or no leisure-time physical activity and 57 % in the other group. The average BMI was 29.2 kg/m^2^ in those who reported little or no leisure-time physical activity, while 27 % had Obesity Class I and 12 % had Obesity Class II. The average BMI was 28.2 kg/m^2^ in those who reported medium or high leisure-time physical activity, while 22 % had Obesity Class I and 7 % had Obesity Class II. The average waist circumference was 94.5 cm in those who reported little or no leisure-time physical activity and 56 % had a high waist circumference. The average was circumference was 92.6 cm in those who reported medium or high leisure-time physical activity and 42 % had a high value. The proportion with no education was 14.5 % in the group that reported little or no leisure-time physical activity and 6.4 % in the group that reported medium or high leisure-time physical activity. The proportion in the lowest income tertile was 49 % in the group that reported little or no activity and 39 % in the other group. More than 70 % of cohort members were married or with partner. The proportions who smoked, who drank alcohol, and who ate fruit and vegetables frequently were somewhat higher in those who reported medium or high leisure-time physical activity. Around 70 % of cohort members reported sleeping 7-9 hours per night.

**Table 1.** Participants’ characteristics at baseline according to leisure-time physical activity volume*.

|  |  | Leisure-time physical activity volume |  |
| --- | --- | --- | --- |
|  |  | None or low | Medium or high |
| Number (%) |  | 122,648 (80.13) | 30,412 (19.87) |
| Age, years, mean (SD) |  | 52 (13) | 52 (13) |
| Sex |  |  |  |
|  | Male, n (%) | 37,004 (30.17) | 12,995 (42.73) |
|  | Female, n (%) | 85,644 (69.83) | 17,417 (57.27) |
| Body mass index, kg/m<sup>2</sup>, mean (SD) |  | 29.2 (5.2) | 28.2 (4.5) |
| Body mass index category |  |  |  |
|  | Normal, n (%) | 23,173 (18.89) | 6,975 (22.94) |
|  | Underweight, n (%) | 615 (0.50) | 126 (0.41) |
|  | Overweight, n (%) | 51,093 (41.66) | 14,596 (47.99) |
|  | Obesity Class I, n (%) | 32,958 (26.87) | 6,602 (21.71) |
|  | Obesity Class II, n (%) | 14,809 (12.07) | 2,113 (6.95) |
| Waist circumference, cm, mean (SD) |  | 94.5 (11.9) | 92.6 (11.1) |
| Waist circumference category |  |  |  |
|  | Low, n (%) | 53,607 (43.71) | 17,539 (57.67) |
|  | High, n (%) | 69,041 (56.29) | 12,873 (42.33) |

**Table 1 (continued).** Participants' characteristics at baseline according to leisure-time physical activity volume\*
|  |  | Leisure-time physical activity volume |  |
| --- | --- | --- | --- |
|  |  | None or low | Medium or high |
| Education |  |  |  |
|  | None, n (%) | 17,731 (14.46) | 1,948 (6.41) |
|  | Some elementary, n (%) | 60,220 (49.12) | 11,481 (37.76) |
|  | Some high-school, n (%) | 28,759 (23.46) | 8,922 (29.34) |
|  | Some college, n (%) | 7,009 (5.72) | 2,993 (9.84) |
|  | Some university, n (%) | 8,876 (7.24) | 5,060 (16.64) |
| Income |  |  |  |
|  | Tertile 1 (lowest), n (%) | 59,938 (48.87) | 11,976 (39.38) |
|  | Tertile 2, n (%) | 25,505 (20.80) | 5,456 (17.94) |
|  | Tertile 3 (highest), n (%) | 37,205 (30.33) | 12,980 (42.68) |
| Civil status |  |  |  |
|  | Not married or with partner, n (%) | 19,570 (15.96) | 4,949 (16.27) |
|  | Married or with partner, n (%) | 87,825 (71.61) | 22,082 (72.61) |
|  | Widowed, n (%) | 15,247 (12.43) | 3,379 (11.11) |
| Smoking |  |  |  |
|  | Never, n (%) | 61,781 (50.41) | 37,937 (30.95) |
|  | Former, n (%) | 22,839 (18.64) | 7,322 (24.09) |
|  | Current, n (%) | 37,937 (30.95) | 10,049 (33.07) |

**Table 1 (continued).** Participants' characteristics at baseline according to leisure-time physical activity volume\*
|  |  | Leisure-time physical activity volume |  |
| --- | --- | --- | --- |
|  |  | None or low | Medium or high |
| Alcohol drinking |  |  |  |
|  | Never, n (%) | 25,859 (21.09) | 4,465 (14.69) |
|  | Occasionally, n (%) | 89,773 (73.21) | 23,224 (76.39) |
|  | At least once per week, n (%) | 5,639 (4.60) | 2,345 (7.71) |
|  | Daily, n (%) | 1,346 (1.10) | 367 (1.21) |
| Fruit and vegetable intake |  |  |  |
|  | Never, n (%) | 1,443 (1.18) | 180 (0.59) |
|  | One or two days per week, n (SD) | 22,757 (18.56) | 3,111 (10.23) |
|  | Three or four days per week, n (SD) | 35,895 (29.27) | 6,790 (22.33) |
|  | Five or more days per week, n (SD) | 62,526 (50.99) | 20,328 (66.85) |
| Sleep duration |  |  |  |
|  | 7-9 hours/night, n (%) | 82,695 (67.42) | 20,679 (68.00) |
|  | <7 or >9 hours/night, n (%) | 39,953 (32.58) | 9,733 (32.00) |

\*Leisure-time physical activity volume was derived from questions about frequency of sport and exercise (less than weekly; one to two times per week; three or more times per week) and duration of sport and exercise (less than 30 minutes per session; 30-60 minutes per session; more than 60 minutes per session) as described in Supplemental Table S1 in the online supplement.

Table 2 shows hazard ratios for all-cause mortality according to leisure-time physical activity volume. Participants were followed for 17.6 (3.9) years [mean (SD)] and there was a total of 25,317 deaths from all causes during 2,694,706 person-years of follow-up. Compared with the group that reported little or no leisure-time physical activity, the hazard ratio (95 % confidence interval) was 0.82 (0.79, 0.85) in the group that reported medium or high leisure-time physical activity after adjusting for age and sex (Model 1). Hazard ratios were similar after adjusting for age, sex, BMI, and waist circumference (Model 2) and after adjusting for age, sex, education, income, civil status, smoking, alcohol, fruit and vegetable intake, and sleep (Model 3). Table 2 also shows hazard ratios for all-cause mortality according to BMI. Compared with the group with normal BMI, the hazard ratio was 0.98 (0.95, 1.01) in the group with overweight, 1.08 (1.05, 1.13) in the group with Obesity Class I, and 1.46 (1.40, 1.53) in the group with Obesity Class II after adjusting for age and sex (Model 1). Overweight, Obesity Class I, and Obesity Class II were not associated with increased risk of all-cause mortality after adjustment for age, sex, and waist circumference (Model 2). Obesity Class I and Obesity Class II were associated with increased risk of all-cause mortality after adjusting for age, sex, education, income, civil status, smoking, alcohol, fruit and vegetable intake, and sleep (Model 3). Finally, Table 2 shows hazard ratios for all-cause mortality according to waist circumference when abdominal obesity was defined as waist circumference ≥88 cm in women and ≥102 cm in men. Compared with the group with low waist circumference, the hazard ratio was 1.27 (1.23, 1.30) in the group with high waist circumference after adjusting for age and sex (Model 1). High waist circumference was also associated with increased risk of all-cause mortality after adjustment for age, sex, and BMI (Model 2) and after adjusting for age, sex, education, income, civil status, smoking, alcohol, fruit and vegetable intake, and sleep (Model 3). Supplemental Table S2 in the online supplement shows that similar patterns were observed in the sensitivity analysis when abdominal obesity was defined as waist circumference ≥80 cm in women and ≥90 cm in men, although the associations were weaker.

**Table 2.**
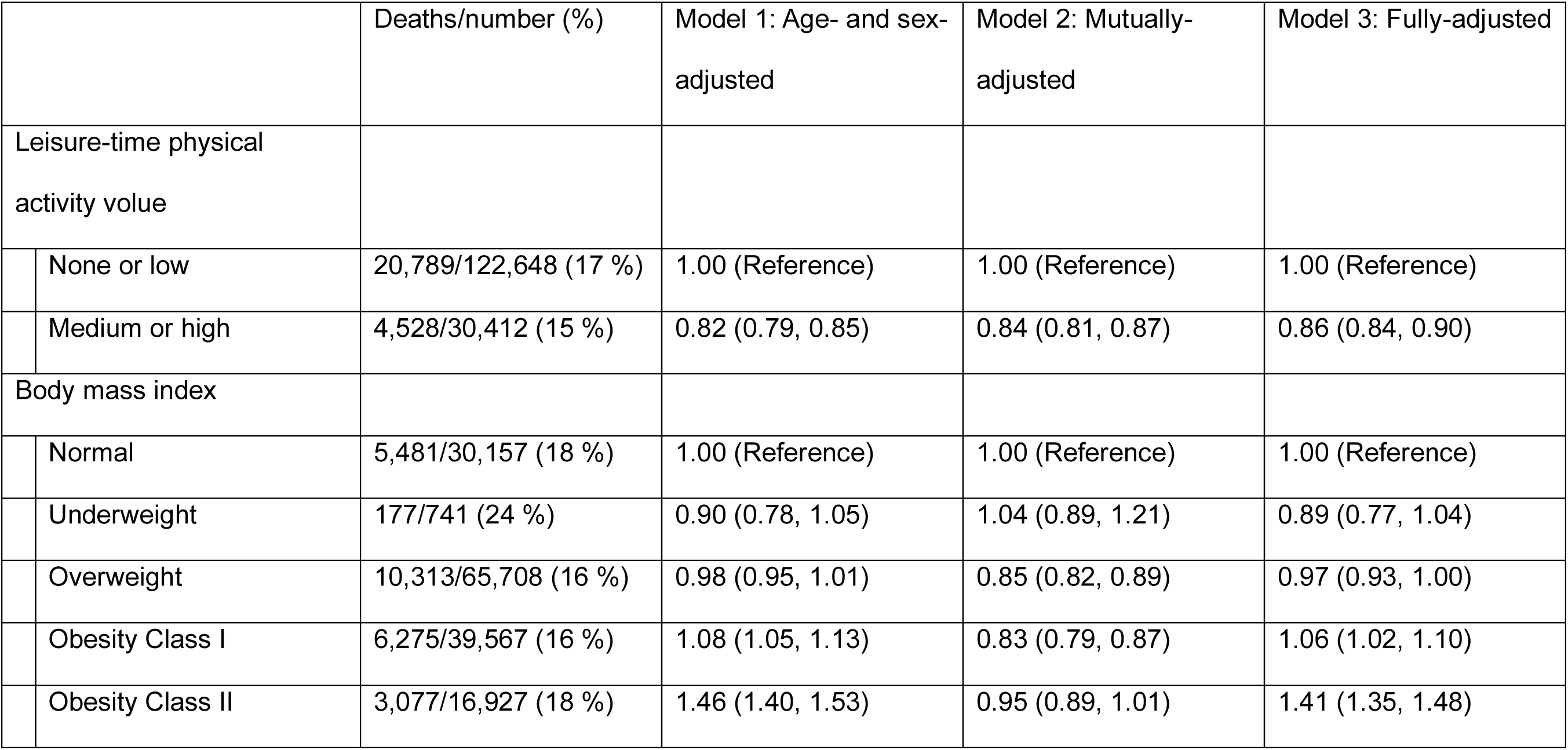
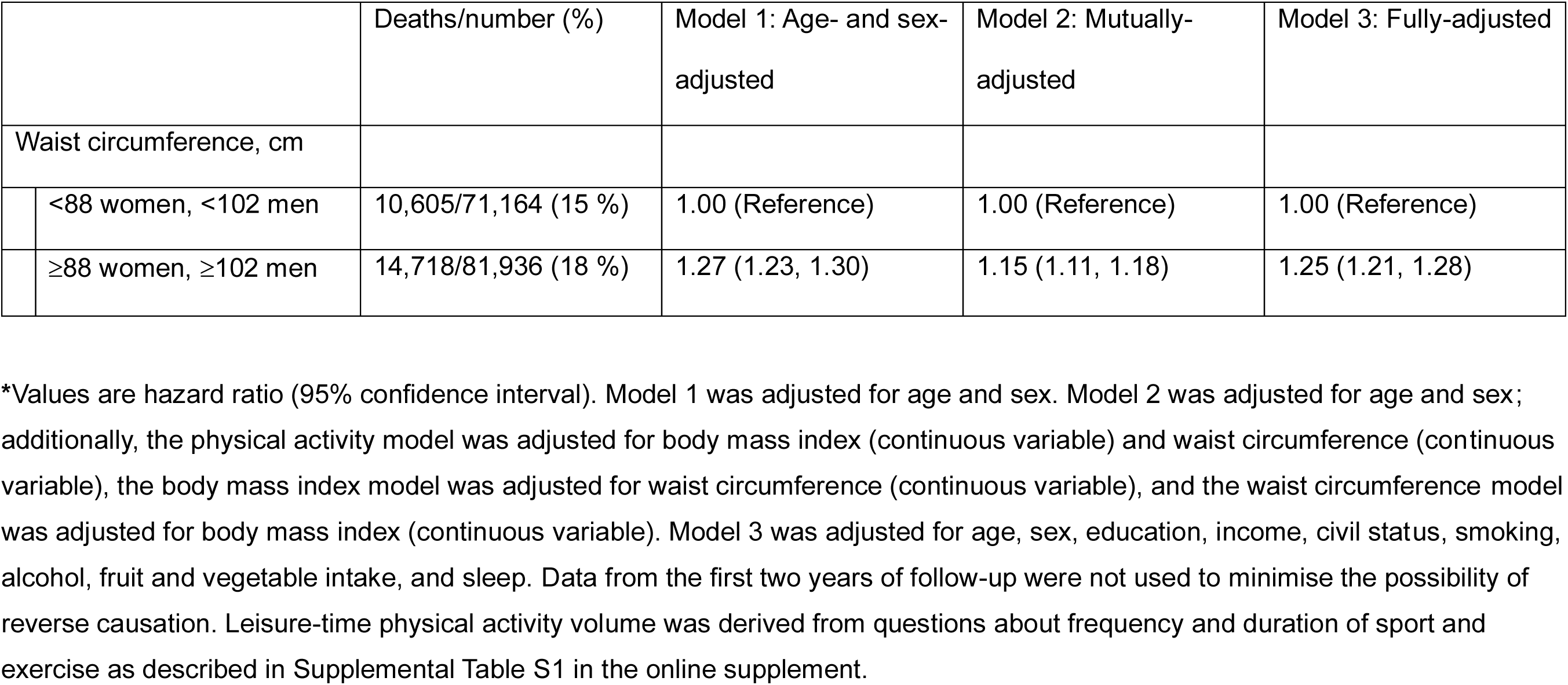
Hazard ratios for all-cause mortality according to leisure-time physical activity volume, body mass index, and waist circumference*.

|  | Deaths/number (%) | Model 1: Age- and sex-adjusted | Model 2: Mutually-adjusted | Model 3: Fully-adjusted |
| --- | --- | --- | --- | --- |
| Leisure-time physical activity volume |  |  |  |  |
| None or low | 20,789/122,648 (17 %) | 1.00 (Reference) | 1.00 (Reference) | 1.00 (Reference) |
| Medium or high | 4,528/30,412 (15 %) | 0.82 (0.79, 0.85) | 0.84 (0.81, 0.87) | 0.86 (0.84, 0.90) |
| Body mass index |  |  |  |  |
| Normal | 5,481/30,157 (18 %) | 1.00 (Reference) | 1.00 (Reference) | 1.00 (Reference) |
| Underweight | 177/741 (24 %) | 0.90 (0.78, 1.05) | 1.04 (0.89, 1.21) | 0.89 (0.77, 1.04) |
| Overweight | 10,313/65,708 (16 %) | 0.98 (0.95, 1.01) | 0.85 (0.82, 0.89) | 0.97 (0.93, 1.00) |
| Obesity Class I | 6,275/39,567 (16 %) | 1.08 (1.05, 1.13) | 0.83 (0.79, 0.87) | 1.06 (1.02, 1.10) |
| Obesity Class II | 3,077/16,927 (18 %) | 1.46 (1.40, 1.53) | 0.95 (0.89, 1.01) | 1.41 (1.35, 1.48) |

**Table 2 (continued).** Hazard ratios for all-cause mortality according to leisure-time physical activity volume, body mass index, and waist circumference\*
|  | Deaths/number (%) | Model 1: Age- and sex-adjusted | Model 2: Mutually-adjusted | Model 3: Fully-adjusted |
| --- | --- | --- | --- | --- |
| Waist circumference, cm |  |  |  |  |
| <88 women, <102 men | 10,605/71,164 (15 %) | 1.00 (Reference) | 1.00 (Reference) | 1.00 (Reference) |
| ≥88 women, ≥102 men | 14,718/81,936 (18 %) | 1.27 (1.23, 1.30) | 1.15 (1.11, 1.18) | 1.25 (1.21, 1.28) |

\*Values are hazard ratio (95% confidence interval). Model 1 was adjusted for age and sex. Model 2 was adjusted for age and sex; additionally, the physical activity model was adjusted for body mass index (continuous variable) and waist circumference (continuous variable), the body mass index model was adjusted for waist circumference (continuous variable), and the waist circumference model was adjusted for body mass index (continuous variable). Model 3 was adjusted for age, sex, education, income, civil status, smoking, alcohol, fruit and vegetable intake, and sleep. Data from the first two years of follow-up were not used to minimise the possibility of reverse causation. Leisure-time physical activity volume was derived from questions about frequency and duration of sport and exercise as described in Supplemental Table S1 in the online supplement.

Table 3 shows hazard ratios for all-cause mortality according to leisure-time physical activity volume and BMI. Obesity Class I and, particularly, Obesity Class II were associated with increased risk in participants who reported little or no activity. For example, compared with the group that reported little or no activity and had a normal BMI, the hazard ratio (95 % confidence interval) was 1.40 (1.33, 1.47) in the group that reported little or no activity and had Obesity Class II after adjustment for potential confounders. Leisure-time physical activity reduced the risk of all-cause mortality associated with obesity. For example, the hazard ratio was 1.18 (1.06, 1.32) in the group that reported medium or high activity and had Obesity Class II after adjustment for potential confounders. Table 3 also shows hazard ratios for CVD mortality according to leisure-time physical activity volume and BMI. Obesity Class I and Obesity Class II were associated with increased risk in participants who reported little or no activity. For example, compared with the group that reported little or no activity and had a normal BMI, the hazard ratio was 1.49 (1.37, 1.63) in the group that reported little or no activity and had Obesity Class II after adjustment for potential confounders. Leisure-time physical activity did not reduce the risk of CVD mortality associated with Obesity Class I but did reduce the risk associated with Obesity Class II: the hazard ratio was 1.24 (1.02, 1.50) in the group that reported medium or high activity and had Obesity Class II after adjustment for potential confounders. Finally, Table 3 shows hazard ratios for cancer mortality according to leisure-time physical activity volume and BMI. Both Obesity Class I and Obesity Class II were associated with considerable risk in participants who reported little or no activity. For example, compared with the group that reported little or no activity and had a normal BMI, the hazard ratio was 1.43 (1.24, 1.64) in the group that reported little or no activity and had Obesity Class II after adjustment for potential confounders. Leisure-time physical activity eliminated the risk of cancer mortality associated with Obesity Class I. Leisure-time physical activity reduced the risk of cancer mortality associated with Obesity Class II: the hazard ratio was 1.34 (1.01, 1.79) in the group that reported medium or high activity and had Obesity Class II after adjustment for potential confounders.

**Table 3.** Hazard ratios for mortality according to leisure-time physical activity volume and body mass index*.

| Leisure-time physical activity volume |  | Body mass index |  |  |  |
| --- | --- | --- | --- | --- | --- |
|  |  | Normal | Overweight | Obesity Class I | Obesity Class II |
| *All-cause mortality* |  |  |  |  |  |
|  | None or low | 1.00 (Reference)<br>[4,450/23,127 (19.2 %)] | 0.96 (0.92, 0.99)<br>[8,186/51,012 (16.0 %)] | 1.04 (1.00, 1.09)<br>[5,236/32,907 (15.9 %)] | 1.40 (1.33, 1.47)<br>[2,708/14,784 (18.3 %)] |
|  | Medium or high | 0.84 (0.78, 0.90)<br>[1,014/6,961 (14.6 %)] | 0.85 (0.81, 0.90)<br>[2,095/14,575 (14.4 %)] | 0.94 (0.88, 1.01)<br>[1,028/6,594 (15.6 %)] | 1.18 (1.06, 1.32)<br>[363/2,111 (17.2 %)] |
| *CVD mortality* |  |  |  |  |  |
|  | None or low | 1.00 (Reference)<br>[1,514/23,127 (6.5 %)] | 1.00 (0.94, 1.07)<br>[2,712/51,012 (5.3 %)] | 1.13 (1.05, 1.22)<br>[1,725/32,907 (5.2 %)] | 1.49 (1.37, 1.63)<br>[874/14,784 (5.9 %)] |
|  | Medium or high | 0.90 (0.79, 1.01)<br>[351/6,961 (5.0 %)] | 0.98 (0.89, 1.07)<br>[744/14,575 (5.1 %)] | 1.12 (1.00, 1.26)<br>[369/6,594 (5.6 %)] | 1.24 (1.02, 1.50)<br>[116/2,111 (5.5 %)] |
| *Cancer mortality* |  |  |  |  |  |
|  | None or low | 1.00 (Reference)<br>[483/23,127 (2.1 %)] | 1.09 (0.98, 1.22)<br>[1,017/51,012 (2.1 %)] | 1.33 (1.19, 1.49)<br>[790/32,907 (2.4 %)] | 1.43 (1.24, 1.64)<br>[352/14,784 (2.4 %)] |
|  | Medium or high | 1.04 (0.87, 1.25)<br>[148/6,961 (2.1 %)] | 1.08 (0.94, 1.25)<br>[316/14,575 (2.2 %)] | 1.01 (0.83, 1.22)<br>[131,6,594 (2.0%)] | 1.34 (1.01, 1.79)<br>[52/2,111 (2.5 %)] |

\*Values are hazard ratio (95% confidence interval) [deaths/number (%)]. For CVD mortality, all other causes of death were treated as competing risks. For cancer mortality, all non-cancer deaths were treated as competing risks. The models were adjusted for age, sex, education, income, civil status, smoking, alcohol, fruit and vegetable intake, and sleep. Data from the first two years of follow-up were

Table 4 shows hazard ratios for all-cause mortality according to leisure-time physical activity volume and waist circumference when abdominal obesity was defined as waist circumference ≥88 cm in women and ≥102 cm in men. High waist circumference was associated with increased risk in participants who reported little or no activity. Compared with the group that reported little or no activity and had low waist circumference, the hazard ratio (95 % confidence interval) was 1.24 (1.21, 1.28) in the group that reported little or no activity and had high waist circumference after adjusting for age, sex, education, income, civil status, smoking, alcohol, fruit and vegetable intake, and sleep. Leisure-time physical activity reduced the risk of all-cause mortality associated with high waist circumference: the hazard ratio was 1.08 (1.03, 1.14) in the group that reported medium or high activity and had high waist circumference after adjustment for potential confounders. Table 4 also shows hazard ratios for CVD mortality according to leisure-time physical activity and waist circumference. High waist circumference was associated with increased risk in participants who reported little or no activity. Compared with the group that reported little or no activity and had low waist circumference, the hazard ratio was 1.28 (1.21, 1.35) after adjusting for potential confounders. Leisure-time physical activity reduced the risk of CVD mortality associated with high waist circumference: the hazard ratio was 1.18 (1.08, 1.28) in the group that reported medium or high activity and had high waist circumference after adjustment for potential confounders. Finally, Table 4 shows hazard ratios for cancer mortality according to leisure-time physical activity and waist circumference. High waist circumference was associated with increased risk in participants who reported little or no activity. Compared with the group that reported little or no activity and had low waist circumference, the hazard ratio was 1.26 (1.16, 1.37) after adjusting for potential confounders. Leisure-time physical activity reduced the risk of cancer mortality associated with high waist circumference: the hazard ratio was 1.14 (1.00, 1.30) in the group that reported medium or high activity and had high waist circumference after adjustment for potential confounders. Supplemental Table S3 in the online supplement shows that similar patterns were observed in the sensitivity analysis when abdominal obesity was defined as waist circumference ≥80 cm in women and ≥90 cm in men: high waist circumference was associated with increased risk of all-cause, CVD, and cancer mortality in participants who reported little or no activity and leisure-time physical activity reduced the risks associated with high waist circumference.

**Table 4.** Hazard ratios for mortality according to leisure-time physical activity volume and waist circumference*.

| Leisure-time physical activity volume |  | Low waist circumference:<br><88 cm in women and <102 cm in men | High waist circumference:<br>≥88 cm in women and ≥102 cm in men |
| --- | --- | --- | --- |
| *All-cause mortality* |  |  |  |
|  | None or low | 1.00 (Reference)<br>[8,294/53,526 (15.5 %)] | 1.24 (1.21, 1.28)<br>[12,438/68,919 (18.1 %)] |
|  | Medium or high | 0.88 (0.84, 0.92)<br>[2,283/17,511 (13.0 %)] | 1.08 (1.03, 1.14)<br>[2,240/12,856 (17.4 %)] |
| *CVD mortality* |  |  |  |
|  | None or low | 1.00 (Reference)<br>[2,651/53,526 (5.0 %)] | 1.28 (1.21, 1.35)<br>[4,227/68,919 (6.1 %)] |
|  | Medium or high | 0.97 (0.90, 1.06)<br>[788/17,511 (4.5 %)] | 1.18 (1.08, 1.28)<br>[801/12,856 (6.2%)] |
| *Cancer mortality* |  |  |  |
|  | None or low | 1.00 (Reference)<br>[1,047/53,526 (2.0 %)] | 1.26 (1.16, 1.37)<br>[1,663/68,919 (2.4 %)] |
|  | Medium or high | 0.97 (0.86, 1.10)<br>[337/17,551 (2.0 %)] | 1.14 (1.00, 1.30)<br>[313/12,856 (2.4 %)] |

\*Values are hazard ratio (95% confidence interval) [deaths/number (%)]. For CVD mortality, all other causes of death were treated as competing risks. For cancer mortality, all non-cancer deaths were treated as competing risks. The models were adjusted for age, sex, education, income, civil status, smoking, alcohol, fruit and vegetable intake, and sleep. Data from the first two years of follow-up were not used to minimise the possibility of reverse causation. Leisure-time physical activity volume was derived from questions about frequency and duration of sport and exercise as described in Supplemental Table S1 in the online supplement.

## Discussion

The main objective of this study was to investigate whether leisure-time physical activity reduced the risks of mortality associated with general and abdominal obesity in adults in Mexico. We found that high BMI was associated with increased risk of mortality and that leisure-time physical activity reduced the risk of all-cause mortality associated with Obesity Class II, the risk of CVD mortality associated with Obesity Class II, and the risk of cancer mortality associated with Obesity Class I and Obesity Class II. We also found that high waist circumference was associated with increased risk of mortality and that leisure-time physical activity reduced the risks of all-cause, CVD, and cancer mortality associated with high waist circumference. To the best of our knowledge, this is the first cohort study to show that leisure-time physical activity reduces the risks of mortality associated with general and abdominal obesity in Mexico.

Cohort studies in Europe and North America suggest that physical activity reduces the risks of mortality associated with general and abdominal obesity.^8–10^ For example, Sanchez-Lastra and colleagues investigated 295,917 middle-aged men and women from the UK and found that physical activity reduced the risks of mortality associated with elevated BMI or elevated waist circumference.^8^ Tarp and colleagues investigated 34,492 middle-aged men and women in Norway, Sweden, the UK, and the US and also found that physical activity reduced the risks of mortality associated with elevated BMI or elevated waist circumference.^10^ There is some evidence that physical activity also reduces the risks of mortality associated with obesity in Latin America, albeit using pooled data from Latin America and other regions.^28^ Lear and colleagues combined data from 130,845 middle-aged men and women from 17 countries including Argentina, Brazil, Chile, and Colombia in Latin America.^28^ They found that medium and high volumes of physical activity were associated with lower risk of all-cause mortality both in participants with BMI <25 kg/m^2^ and in participants with BMI ≥25 kg/m^2^.^28^ They also found that physical activity was associated with reduced risk of all-cause mortality both in those with normal waist-to-hip ratio and in those with high waist-to-hip ratio.^28^ To the best of our knowledge, the Mexico City Prospective Study is the largest cohort study in Mexico and the present investigation is the first analysis of the combined associations of physical activity and obesity with mortality in Mexico.

The present investigation has important implications for policy and practice. The prevalence of Obesity Class I and, particularly, Obesity Class II is rising every year in men and women in Mexico.^4^ The prevalence of abdominal obesity is also rising every year in adults in Mexico.^4^ The doses of sport and exercise reported in the present study are broadly equivalent to the recommended doses for weight management of 150-300 minutes of leisure-time physical activity per week.^29^ There are many opportunities to take part in such doses of physical activity in Mexico City. The city is home to a weekly physical activity intervention known as *Muévete en Bici* (or, Get on your Bike!).^30^ Every Sunday, more than 60 kilometers of roads are closed to motor vehicles and the streets fill with aound 95,000 walkers, runners, and cyclists.^30^ Approximately 90% of participants in the intervention report taking part in at least 150 minutes of moderate to vigorous physical activity.^31^ One could accumulate another 150 minutes of physical activty across the week by taking advantage of the city’s bike hire scheme, by using the city’s network of cycle lanes and renovated sidewalks, and by visiting the city’s parks, sports centres, and open-air gyms. These policies and practicies are implemented in dozens of neighbourhoods in Mexico City, but it is recognised that more must be done to promote physical activity in children and adults in neighbourhoods of lower socioeconomic status.^32, 33^ Many cities in Latin America have adopted similar policies and practices,^34–36^ and it has been argued that policy makers in other regions should implement comparable measures in order to takle the “pandemic of physical inactivity”.^37, 38^

This study has strengths and limitations. The Mexico City Prospective Study is deemed to be broadly representative of the population aged 35 or older by virtue of the large sample size and the high response rate.^15^ The present investigation included more than 150,000 cohort members and analyses were adjusted for major confounders.^8, 24–26^ Cardiovascular disease and cancer mortality were treated as competing risks. Furthermore, data from the first two years of follow-up were not used to minimize the possibility of reverse causation. Physical activity was self-reported, but there is little risk of misclassification of the active and inactive with such a simple questionnaire. It would be preferable to use both questionnaires and accelerometers to assess physical activity in cohort studies because each method has advantages and disadvantages.^39, 40^ For example, questionnaires might best be used to identify types of sport and exercise and accelerometers might best be used to assess volumes of physical activity. The assessment of diet was relatively simple, but relatively sophisticated assessments suggest that the benefits of physical activity are largely independent of the confounding effects of diet.^28^

In conclusion, this large cohort study suggests that leisure-time physical activity reduces the risks of mortality associated with general and abdominal obesity in Latin America. This study has important implications for policy and practice in the region because the promotion of healthy lifestyles may be a more effective strategy than the promotion of weight loss in reducing the burden of obesity at the population level.

## Data Availability

Mexico City Prospective Study data are available for open-access data requests. The data access policy is described online: http://www.ctsu.ox.ac.uk/research/mcps.

## Acknowledgements

We thank everyone who took part in the study. This research has been conducted using Mexico City Prospective Study (MCPS) Data under Application Number 2022-015.

## Funding

The present analysis received no specific funding. The Mexico City Prospective Study has received funding from the Mexican Health Ministry, the National Council of Science and Technology for Mexico, Wellcome, and core grants from the UK Medical Research Council to the MRC Population Health Research Unit at the University of Oxford (https://www.ctsu.ox.ac.uk/research/mcps). The funding sources had no role in the design, conduct, or analysis of the study or in the decision to submit the manuscript for publication.

## Conflicts of interest

None.

## Authors’ contributions

O’Donovan had a role in study conceptualization, data acquisition, formal analysis, methodology, writing the original draft, and editing the drafts. Petermann-Rocha, Ferrari, Medina, Apolinar-Jiménez, and Sarmiento had a role in reviewing and editing. O’Donovan is guarantor.

## Notes

### Competing Interest Statement

The authors have declared no competing interest.

### Author Declarations

The Mexico City Prospective Study was approved by the Mexican Ministry of Health, the Mexican National Council of Science and Technology (approval number 0595 P-M), and the Central Oxford Research Ethics Committee (approval number C99.260).

