## Supplementary material for "Leisure-time physical activity reduces the risks of mortality associated with general and abdominal obesity in adults in Mexico": online supplement

**Supplemental Table S1.** Leisure-time physical activity volume as derived from questions about frequency and duration of sport and exercise*

| Frequency of sport and exercise | Duration of sport and exercise sessions | | |
|---|---|---|---|
| | Less than 30 minutes | 30-60 minutes | More than 60 minutes |
| None | None | None | None |
| Less than weekly | Low | Low | Low |
| Once or twice per week | Low | Medium | Medium |
| Three or more times per week | Medium | High | High |

*Leisure-time physical activity volume was derived from three questions about frequency and duration of sport and exercise and was categorised as none, low, medium, or high. Participants were first asked: “Do you take part in any sport or exercise?” (Yes; No; Don’t know, or no response.) Participants were then asked: “How many times per week do you exercise?” (Less than once per week; One or two times per week; Three or more times per week; Don’t know, or no response.) Finally, participants were asked: “When you exercise, for how many minutes do you do it?” (Less than 30 minutes; From 30 to 60 minutes; More than 60 minutes; Don’t know, or no response.)

**Supplemental Table S2.** Hazard ratios for all-cause mortality according to waist circumference, with abdominal obesity defined as waist circumference ≥80 cm in women and ≥90 cm in men*

| Waist circumference, cm | Deaths/number (%) | Model 1: Age- and sex-adjusted | Model 2: Mutually-adjusted | Model 3: Fully-adjusted |
|---|---|---|---|---|
| <80 women, <90 men | 3,320/24,331 (13.7 %) | 1.00 (Reference) | 1.00 (Reference) | 1.00 (Reference) |
| ≥80 women, ≥90 men | 22,003/128,769 (17.1 %) | 1.17 (1.13, 1.21) | 1.02 (0.98, 1.06) | 1.15 (1.11, 1.20) |

*Values are hazard ratio (95% confidence interval) [deaths/number (%)]. Model 1 was adjusted for age and sex. Model 2 was adjusted for age, sex, and body mass index (continuous variable). Model 3 was adjusted for age, sex, education, income, civil status, smoking, alcohol, fruit and vegetable intake, and sleep. Data from the first two years of follow-up were not used to minimise the possibility of reverse causation. Abdominal obesity was defined as waist circumference ≥80 cm in women and ≥90 cm in men in this sensitivity analysis because of the low stature of adults in Mexico.

**Supplemental Table S3.** Hazard ratios for mortality according to leisure-time physical activity volume and waist circumference, with abdominal obesity defined as waist circumference ≥80 cm in women and ≥90 cm in men*

| Leisure-time physical activity volume | | Low waist circumference: <80 cm in women and <90 cm in men | High waist circumference: ≥80 cm in women and ≥90 cm in men |
|---|---|---|---|
| *All-cause mortality* | | | |
| | None or low | 1.00 (Reference) [2,590/17,801 (14.6 %)] | 1.14 (1.09, 1.19) [18,142/104,644 (17.3 %)] |
| | Medium or high | 0.85 (0.78, 0.92) [724/6,489 (11.2 %)] | 0.99 (0.94, 1.04) [3,799/23,878 (15.9 %)] |
| *CVD mortality* | | | |
| | None or low | 1.00 (Reference) [811/17,801 (4.6 %)] | 1.18 (1.09, 1.28) [6,067/104,644 (5.8 %)] |
| | Medium or high | 0.96 (0.83, 1.11) [246/6,489 (3.8 %)] | 1.11 (1.01, 1.21) [1,343/23,878 (5.6 %)] |
| *Cancer mortality* | | | |
| | None or low | 1.00 (Reference) [314/17,891 (1.8 %)] | 1.22 (1.08, 1.37) [2,396/104,644 (2.3 %)] |
| | Medium or high | 0.98 (0.79, 1.22) [110/6,489 (1.7 %)] | 1.12 (0.97, 1.29) [540/23,878 (2.3 %)] |

*Values are hazard ratio (95% confidence interval) [deaths/number (%)]. For CVD mortality, all other causes of death were treated as competing risks. For cancer mortality, all non-cancer deaths were treated as competing risks. The models were adjusted for age, sex, education, income, civil status, smoking, alcohol, fruit and vegetable intake, and sleep. Data from the first two years of follow-up were not used to minimise the possibility of reverse causation. Leisure-time physical activity volume was derived from questions about frequency and duration of sport and exercise as described in Supplemental Table 1 in the online supplement. Abdominal obesity was defined as waist circumference ≥80 cm in women and ≥90 cm in men in this sensitivity analysis because of the low stature of adults in Mexico.
